# ASSOCIATION BETWEEN PERIODONTITIS AND METABOLIC SYNDROME IN A FAMILY HEALTH UNIT IN SALVADOR-BA

**DOI:** 10.1101/2021.06.03.21258301

**Authors:** Julita Maria Freitas Coelho, Glaúcia Alencar Ponte, Isaac Suzart Gomes-Filho, Johelle de Santana Passos, Simone Seixas da Cruz, Ana Claudia Morais Godoy Figueiredo, Sarah dos Santos Conceição, Roberta Borges Silva, Magno Conceição das Mêrces

**Affiliations:** Doutora em Saúde Coletiva, Instituto Federal de Educação da Bahia; Mestre em Saúde Coletiva, Secretaria Municipal de Saúde de Salvador-Ba, Brasil; Pós Doutor em Saúde Coletiva, Universidade Estadual de Feira de Santana; Doutora em Saúde Coletiva, Universidade Federal da Bahia; Pós Doutora em Saúde Coletiva, Universidade Federal do Recôncavo da Bahia; Doutora em Saúde Coletiva, Universidade de Brasília; Doutoranda em Saúde Coletiva, Universidade de Brasília; Mestre em Nutrição Humana, Ministério da Saúde, Brasil; Doutor em Ciências da Saúde, Universidade do Estado da Bahia, Brasil

**Keywords:** Metabolic Syndrome, Periodontal Disease, Periodontitis, Epidemiology

## Abstract

**Introduction:** The metabolic syndrome is characterized by multiple disorders, and the in periodontitis, inflammation occurs in the tissues supporting the tooth, where in this process it is believed that the migration of oral bacteria and byproducts to the circulatory system occurs, with a systemic spread of inflammatory mediators. This study aims to verify the effect of periodontitis on the occurrence of the metabolic syndrome.

**Method:** Cross-sectional study with 90 users of the Family Health Unit from Vale do Cambonas in Salvador-BA. Questionnaires were applied, physical / anthropometric and dental evaluation were performed, record of the results of laboratory tests was registered and evaluation of medical records.

**Data analysis:** A bivariate and stratified analysis was performed, obtaining means and standard deviation for continuous variables, absolute and relative frequency for all variables, and multiple conditional logistic regression was performed to obtain the final model adjusted for potential confounders.

**Results:** In the final sample 46,67% of participants had metabolic syndrome according NECP-ATP III criterion and 30, 00% had periodontitis. From those, 20.00% had severe periodontitis, 10% moderate periodontitis and none with mild periodontitis, according to the criterion proposed by Gomes-Filho et al. (2018). There was statistical significance in the association between periodontitis and MetS (_ORcrude_ = 2.58, 95% CI [1.02 - 6.55]) / (_ORadjusted_ = 2.63, 95% CI [1.01 - 6.80]) and severe periodontitis and MetS (_ORcrude_ = 3.86, 95% CI [1.24 - 11.98]) / (_ORadjusted_ = 4.14, 95% CI [1.29-13.29]).

**Conclusion:** The main findings of this study indicate a positive association between periodontitis and metabolic syndrome, with a higher effect when the exposure was severe periodontitis.

## INTRODUCTION

Metabolic syndrome (MetS) is defined as a complex disorder, represented by metabolic changes, such as dyslipidemia, arterial hypertension, glucose intolerance, central obesity and insulin resistance, which commonly occur together^1–3^. People diagnosed with MetS have an increased risk for cardiovascular adverse outcomes^4^, such as death caused by coronary disease ^5–8^, and higher risk for type 2 diabetes^9^.

Thus, World Health Organization warns in regards to the increasing of non-communicable diseases (NCD) that compose MetS and for the need of more studies in order to inform necessary control measures^10^. Recent studies showed the evolution of MetS, with a prevalence varying from 25% in Middle Eastern countries^11^ to 50.2% in India^12^. In the United States of America (USA), the prevalence of MetS was estimated at 32.2% in the overall population, which increased for 34.6% in individuals older than 70 years old^13^.

In Brazil, in spite of the lack of studies, a high prevalence of MetS has been reported^14^. Specifically, on a population based study by Oliveira et al. (2020)^15^, that estimated a mean prevalence of 38.4%; from these, 16.7% affects people at 18 to 39 years old, 45.7% happens with those aged 40 to 59 years old and 66.1% in those aged 60 years old or above. A higher prevalence was observed on the indigenous population, 63.4%^16^. Moreover, the prevalence of MetS in Brazil is higher in women, with a lower educational level and in the elderly^15^.

Studies show that a chronic inflammatory process may predispose MetS due to the systemic dissemination of immunologic mediators and the increase in the production of inflammatory proteins, such as reactive C protein^17–21^. Besides, systemic inflammation can cause insulin resistance (IR) and/or hyperglycemia^19^.

Therefore, periodontitis acts as a precursor of systemic inflammatory processes, since there is gingival inflammation associated with damage of periodontal ligament and alveolar bone, and also with the contamination of root cement, which is associated to the presence of specific gram-negative anaerobic bacteria ^22–24^. It is believed that there is a migration of oral bacteria and byproducts directed to the blood flow, with a systemic dissemination of local inflammatory mediators. Therefore, periodontitis may be able to stimulate^17^ inflammatory systemic responses and to contribute with MetS^25–30^.

Studies show a positive association between periodontitis and metabolic syndrome ^20,26,27,31–33^, varying according to population groups. Thus, the aim of the present study is to investigate the association between periodontitis and metabolic syndrome in patients of a Family Health Unit (FHU) in Vale do Cambonas, in the county of Salvador, Bahia.

## METHODS

Analytical cross sectional study, with a convenience sample comprised by patients registered in Vale do Cambonas FHU. Inclusion criteria were: ≥ 30 years old; ≥ 4 teeth; and having registry of laboratory results of triglycerides, HDL cholesterol and fasting glucose, dated 180 days before clinical and oral examination. Up to the reporting of the present study, we included 90 participants, and analyses were obtained with a study power of 51%, confidence level of 95% and exposure/non exposure ratio of 1:3, with a periodontits frequency of 40% for the non-exposed group and 20% for the exposed group^34^. Thus, the number of exposed participants was 27 (with periodontitis) and non-exposed was 63 (without periodontitis).

We excluded from our sample participants who have had periodontal treatment in the last three months, with systemic infections, with HIV/AIDS, and pregnant women. Data collection was performed through a questionnaire, physical/anthropometric and dental assessment, and through the analysis of laboratorial examination results and patients registries. The questionnaire comprised personal and sociodemographic data, human biology, health care and dental assistance. Physical and anthropometric examination measured blood pressure in a calm environment, after at least 5 minutes of resting, and with a previous explanation of the procedure. The participant was guided to stay seated in a comfortable position, keeping legs uncrossed and feet on the ground, leaning the back on the chair (V-DBHA, 2007). We used a stethoscope from Premium® and sphygmomanometer from P.A.MED®, both properly calibrated.

Weight and height were measured using a digital (Indi Peso Instrumentos®), with participants barefooted. Weight and height measures comprised Body Mass Index (BMI), according to Associação Brasileira para o Estudo da Obesidade^35^. We measured waist circumference based on the narrowest part of the waist (the space between the lower costal border and the iliac crest), and hip circumference based on the wider part of the hip and bigger lump of the gluteal region (150cm – scale 0.5cm; and through the waist/hip ratio-WHR – waist circumference divided by hip circumference)^7,35^.

Clinical dental evaluation was performed by a dental surgeon, previously trained, in the dental Office of FHU Vale do Cambonas. Periodontal examination was performed using a Williams probe (from Hu-Friedy®), by the measurement of probe depth, given by the distance between gingival margin and the most apical portion of probe penetration, at six sites per tooth (mesial buccal, mid-buccal, distobuccal, mesio-lingual, mid-lingual, and disto-lingual) In these sites we also verified the presence of bleeding upon probing and measured gingival recession, which comprises the distance between gingival margin and cemento-enamel junction. Dental plaque index was also evaluated for the vestibular, lingual, mesial and distal faces from dental units, and we calculated clinical attachment loss (sum between probe depth and recession measure); these procedures were both performed on the same day

Laboratory examinations (triglycerides, fasting glucose, total cholesterol and fractions) were taken by Hospital Sao Rafael (Secretaria Municipal de Saúde de Salvador). The results were obtained from patient’s registries or from the examination reports held by the participants and registered in a clinical Record. Participants that were using antihypertensive or lipid-lowering medication, or with a previous diabetes diagnosis also filled up information regarding blood pressure, triglycerides and fasting glucose, respectively^7^.

The classification by Gomes-Filho et al. (2018)^36^ and Page and Eke (2007)^37^ and Eke (2012)^38^ criterion guided the periodontitis diagnosis and its severity levels, such as severe, moderate and mild periodontitis, and the absence of periodontitis. Standardized criterion by NECP-ATP III^39^ guided metabolic syndrome diagnosis, based on three or more components described in the Box below, which characterize the presence of the disease; International Diabetes Federation (IDF, 2006)^40^ criterion also informed its diagnosis through abdominal circumference increase plus at least two other risk factors, as described below:

### Box 1

Metabolic Syndrome diagnosis criterion (NCEP-ATP III*^39^, IDF**^40^)

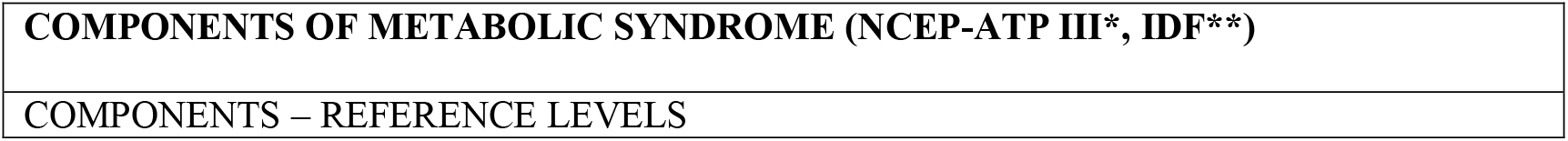
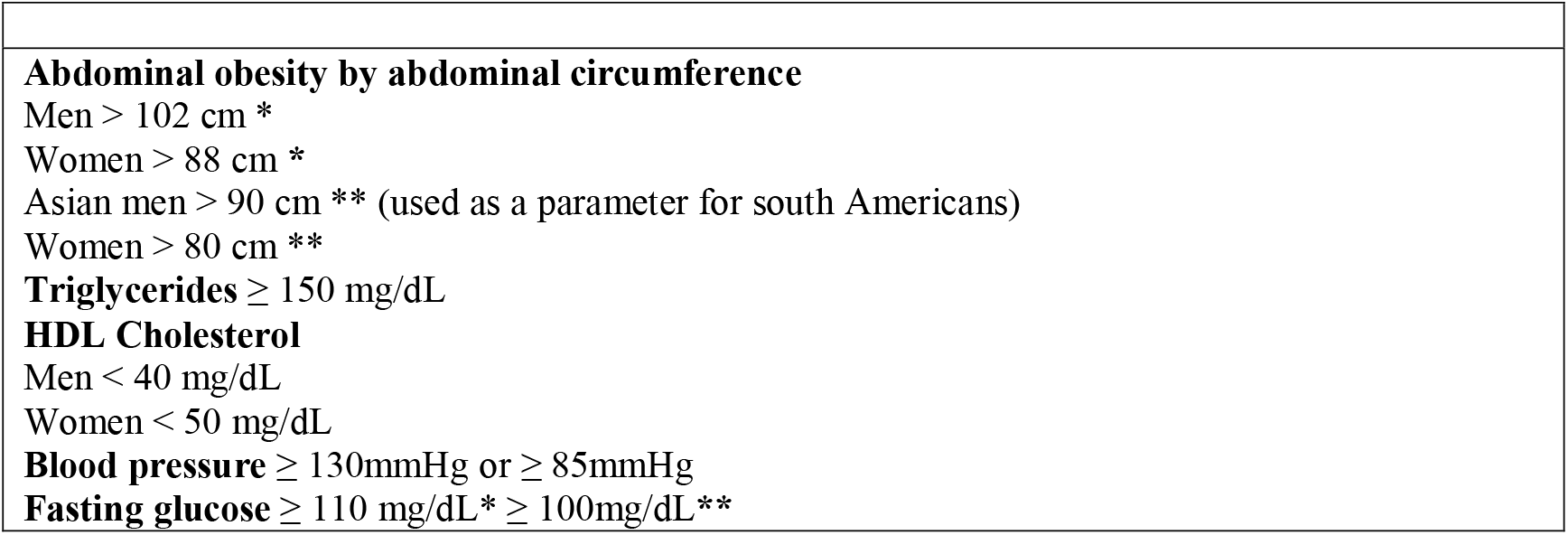

We performed a descriptive analysis for sociodemographic, lifestyle, human biology, health care and dental assistance information. Absolute and relative frequencies from relevant variables were obtained. Bivariate, stratified and multivariate analyses estimated the association between exposure and outcome variables. Pearson Chi-squared and T test estimated measures for independent variables. We adopted a 5% level of significance (p<0,05) and estimated Odds Ratios (OR) and its 95% Confidence Intervals (CI 95%).

Regarding the association between periodontitis and MetS, we evaluated the change on the effect and confounding measures, and we assumed a predictive model using Mantel-Haenszel test, with a significance level of p≤ 0,20. Empirical and theoretical basis guided the selection of potential confounding variables, considering relative differences for each covariable’s adjusted measures and crude association measure above 20%,

Logistic regression was performed by a multivariate analysis, with effect modifying variables verified by maximum likelihood ratio (p<0,05) Since we did not identify any effect change, counfounding analysis was performed by the backward strategy. We considered potential confounding variables as those that promoted a change lower than 10% on the association measure, and we obtained a final model of association between periodontitis and MetS and a final model between severe periodontitis and MetS, both adjusted by age, alcoholic beverage drinking, family income and physical activity.

The adequacy of the regression model was evaluated by the goodness fit of the model, and Hosmer-Lemeshow test was used to evaluate the goodness fit for each model. Odds Ratios (OR) were estimated for: periodontitis (Gomes-Filho et al., 2018)^36^ *vs* MetS (NECP-ATP III)^39^, severe periodontitis (Gomes-Filho et al., 2018)^36^ *vs* MetS (NECP-ATP III)^39^, periodontitis (Gomes-Filho et al., 2018)^36^ *vs* MetS (IDF)^40^, severe periodontitis (Gomes-Filho et al., 2018) ^36^ *vs* MetS (IDF)^40^, periodontitis (Page e Eke, 2007;Eke et al., 2012)^37,38^ *vs* MetS (NECP-ATP III)^39^, severe periodontitis (Page e Eke, 2007;Eke et al., 2012)^37,38^ *vs* MetS (NECP-ATP III)^39^, periodontitis (Page e Eke, 2007;Eke et al., 2012)^37,38^ *vs* MetS (IDF)^40^ e severe periodontitis (Page e Eke, 2007;Eke et al., 2012)^37,38^ *vs* MetS (IDF)^40^. Statistical analyses were performed on STATA 11.

## RESULTS

Ninety participants, including 57 women, comprised this study sample. Individuals diagnosed with MetS were, on average, 54±12 years old, ranging from 30 to 77 years old. There was a statistical significant difference for guidance to oral health (p= 0,02) regarding lifestyle and oral health, according to MetS diagnosis based on NCEP-ATP III^39^ criterion (TABLE 1).

**Table 1.** Sociodemographic and lifestyle characteristics according to NCEP- ATP III criterion for Metabolic Syndrome diagnosis. Salvador, BA, Brazil, 2018. (n= 90)

| Characteristics | MetS Diagnosis (NCEP- ATPIII) |  |  |  | p* |
| --- | --- | --- | --- | --- | --- |
|  | Yes<br>(n= 42) |  | No<br>(n = 48) |  |  |
| Age (years) | n | % | n | % |  |
| < 59 | 22 | 52.38 | 26 | 54.17 |  |
| ≥ 59 | 20 | 47.62 | 22 | 45.83 |  |
| Mean±sd | 54±12 |  | 52±11 |  | 0.87 |
| Median | 55 |  | 52 |  |  |
| Mínimum-Máximo | 30-75 |  | 30-77 |  |  |
| Gender |  |  |  |  |  |
| Female | 30 | 63.64 | 27 | 47.37 | 0.19 |
| Male | 12 | 36.36 | 21 | 52.63 |  |
| Race/skin color |  |  |  |  |  |
| White/yellow | 1 | 2.38 | 5 | 10.42 | 0.13 |
| Black/Brown/other | 41 | 97.62 | 43 | 89.58 |  |
| Marital status |  |  |  |  |  |
| Married/other type | 22 | 52.38 | 24 | 50.00 | 0.82 |
| Single/divorced/widowed | 20 | 47.62 | 24 | 50.00 |  |
| Educational level (years of education) |  |  |  |  |  |
| >4 years | 17 | 40.48 | 18 | 37.5 | 0.77 |
| ≤ 4 years | 25 | 59.52 | 30 | 62.50 |  |
| Family income<sup>1</sup> |  |  |  |  |  |
| >1 minimum wage | 15 | 35.71 | 23 | 47.92 | 0.24 |
| ≤1 minimum wage | 27 | 64.29 | 25 | 52.08 |  |
| Household density<br>(number of people per household) |  |  |  |  |  |
| ≤ 3 people | 28 | 66.67 | 38 | 79.17 | 0.18 |
| >3 people | 14 | 33.33 | 10 | 20.83 |  |
| Number of children |  |  |  |  |  |
| ≤ 1child | 13 | 30.95 | 12 | 25.00 | 0.53 |
| ≥ 2 children | 29 | 69.05 | 36 | 75.00 |  |
| Current smoking habit |  |  |  |  |  |
| Yes | 1 | 2.38 | 5 | 10.42 | 0.21 |
| No | 41 | 97.62 | 43 | 89.58 |  |
| Alcoholic beverage drinking |  |  |  |  |  |
| Yes | 15 | 35.71 | 14 | 29.17 | 0.51 |
| No | 27 | 64.29 | 34 | 70.83 |  |
| Physical activity |  |  |  |  |  |
| Yes | 18 | 42.86 | 20 | 41.67 | 0.91 |
| No | 24 | 57.14 | 28 | 58.33 |  |
| Dental surgeon consultation |  |  |  |  |  |
| ≥ 1 consultation per year | 34 | 80.95 | 36 | 75.00 | 0.50 |
| <1 consultation per year | 8 | 19.05 | 12 | 25.00 |  |
| Guided regarding oral health |  |  |  |  |  |
| Yes | 35 | 83.33 | 29 | 6.42 | 0.02 |
| No | 7 | 16.67 | 19 | 39.68 |  |
| Tooth brushing frequency |  |  |  |  |  |
| < 3x/day | 23 | 47.92 | 25 | 52.08 | 0.80 |
| ≥ 3x/day | 19 | 45.24 | 23 | 54.76 |  |
| Dental flossing |  |  |  |  |  |
| Yes | 21 | 50.00 | 17 | 35.42 | 0.16 |
| No/sometimes | 21 | 50.00 | 31 | 64.58 |  |
| Periodontal treatment |  |  |  |  |  |
| Yes | 12 | 28.57 | 9 | 18.75 | 0.27 |
| No | 30 | 71.43 | 39 | 81.25 |  |

\* Statistical significance level: $p \leq 0.05$ .
<sup>1</sup> Minimum wage value of R\$ 954.00 on data collection date.

According to Gomes-Filho et al. (2018)^36^ diagnosis criterion, 30% of participants had periodontitis, and, from those, 40.48% had MetS; 20% had severe periodontitis, and 30.95% from those had MetS; and 10% had moderate periodontites, 9.52% with MetS.

According to MetS diagnosis, regarding general health status and periodontitis severity, the following variables were statistically different: hypertension (p<0,01), diabetes (p=<0,01), BMI (p =<0,01), waist circumference (p=0,01), waist/hip ratio (p<0,01) and periodontitis diagnosis GOMES-FILHO et al. (2018)^36^ (p= 0,04) (TABLE 2).

**Table 2.**
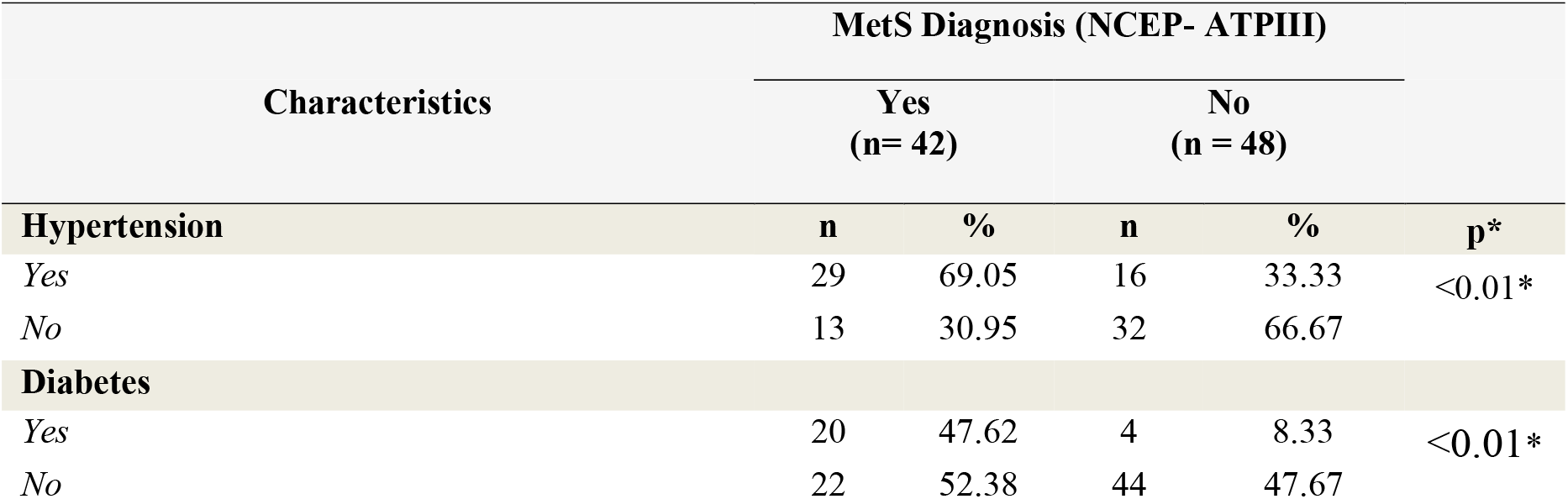
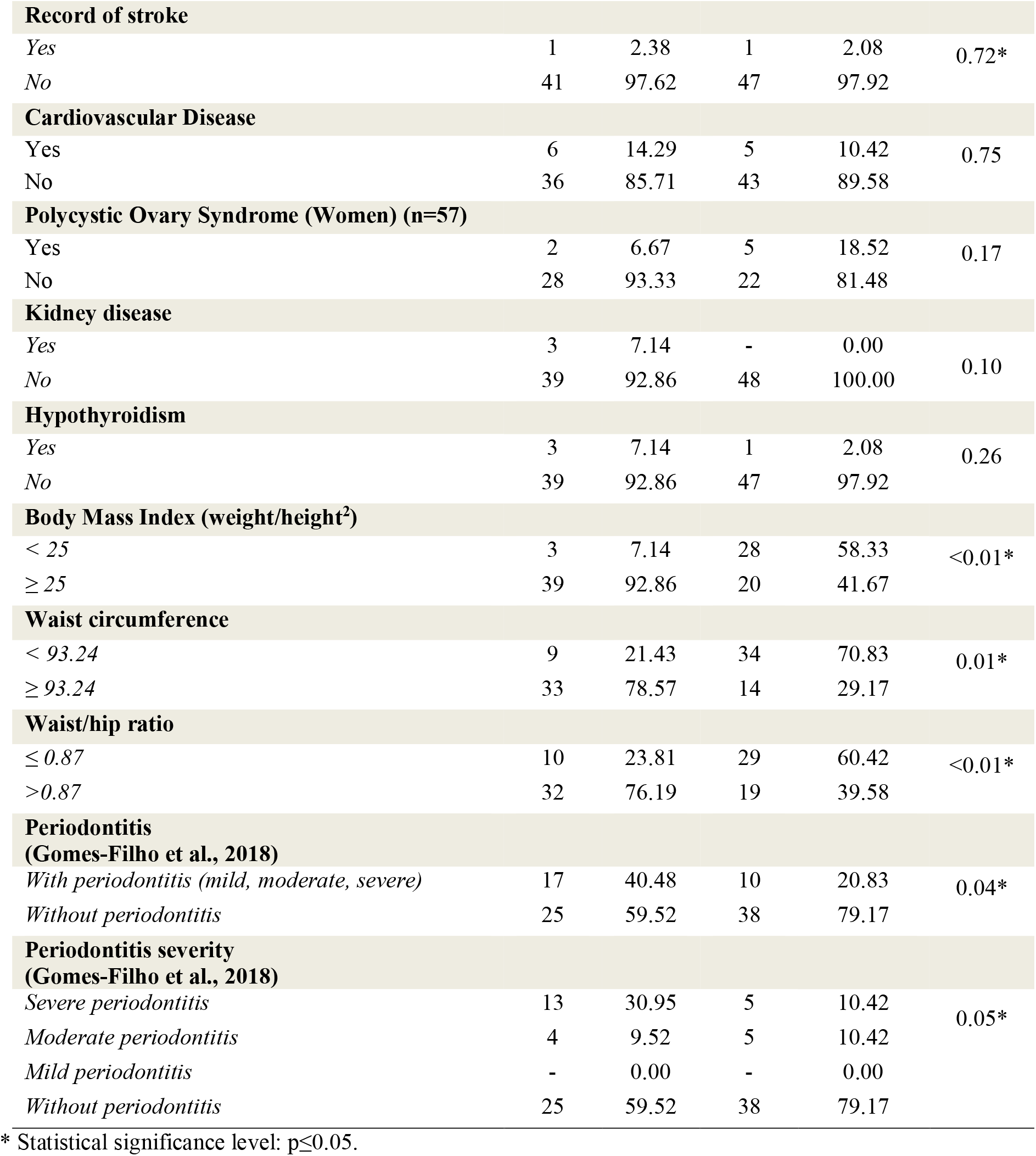
Distribution of characteristics related with general health status and periodontitis severity according to NCEP-ATP III diagnosis criterion for MetS. Salvador, Bahia, Brazil, 2018. (n= 90)

| Characteristics | MetS Diagnosis (NCEP- ATPIII) |  |  |  |  |
| --- | --- | --- | --- | --- | --- |
|  | Yes<br>(n= 42) |  | No<br>(n = 48) |  | p* |
| **Hypertension** | **n** | **%** | **n** | **%** |  |
| *Yes* | 29 | 69.05 | 16 | 33.33 | <0.01* |
| *No* | 13 | 30.95 | 32 | 66.67 |  |
| **Diabetes** |  |  |  |  |  |
| *Yes* | 20 | 47.62 | 4 | 8.33 | <0.01* |
| *No* | 22 | 52.38 | 44 | 47.67 |  |
| Record of stroke |  |  |  |  |  |
| Yes | 1 | 2.38 | 1 | 2.08 | 0.72* |
| No | 41 | 97.62 | 47 | 97.92 |  |
| Cardiovascular Disease |  |  |  |  |  |
| Yes | 6 | 14.29 | 5 | 10.42 | 0.75 |
| No | 36 | 85.71 | 43 | 89.58 |  |
| Polycystic Ovary Syndrome (Women) (n=57) |  |  |  |  |  |
| Yes | 2 | 6.67 | 5 | 18.52 | 0.17 |
| No | 28 | 93.33 | 22 | 81.48 |  |
| Kidney disease |  |  |  |  |  |
| Yes | 3 | 7.14 | - | 0.00 | 0.10 |
| No | 39 | 92.86 | 48 | 100.00 |  |
| Hypothyroidism |  |  |  |  |  |
| Yes | 3 | 7.14 | 1 | 2.08 | 0.26 |
| No | 39 | 92.86 | 47 | 97.92 |  |
| Body Mass Index (weight/height<sup>2</sup>) |  |  |  |  |  |
| < 25 | 3 | 7.14 | 28 | 58.33 | <0.01* |
| ≥ 25 | 39 | 92.86 | 20 | 41.67 |  |
| Waist circumference |  |  |  |  |  |
| < 93.24 | 9 | 21.43 | 34 | 70.83 | 0.01* |
| ≥ 93.24 | 33 | 78.57 | 14 | 29.17 |  |
| Waist/hip ratio |  |  |  |  |  |
| ≤ 0.87 | 10 | 23.81 | 29 | 60.42 | <0.01* |
| >0.87 | 32 | 76.19 | 19 | 39.58 |  |
| Periodontitis<br>(Gomes-Filho et al., 2018) |  |  |  |  |  |
| With periodontitis (mild, moderate, severe) | 17 | 40.48 | 10 | 20.83 | 0.04* |
| Without periodontitis | 25 | 59.52 | 38 | 79.17 |  |
| Periodontitis severity<br>(Gomes-Filho et al., 2018) |  |  |  |  |  |
| Severe periodontitis | 13 | 30.95 | 5 | 10.42 | 0.05* |
| Moderate periodontitis | 4 | 9.52 | 5 | 10.42 |  |
| Mild periodontitis | - | 0.00 | - | 0.00 |  |
| Without periodontitis | 25 | 59.52 | 38 | 79.17 |  |

\* Statistical significance level: $p \leq 0.05$ .

There were statistical significant differences among participants with and without periodontitis regarding the percentual of visible plaque, probe depth, level of clinical attachment, bleeding upon probe and triglycerides, with better results for those without periodontitis (TABLE 3)

**Table 3.**
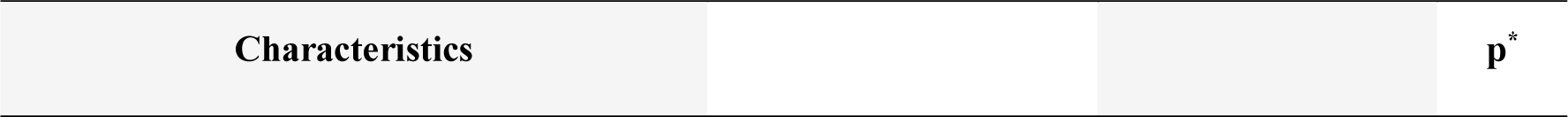
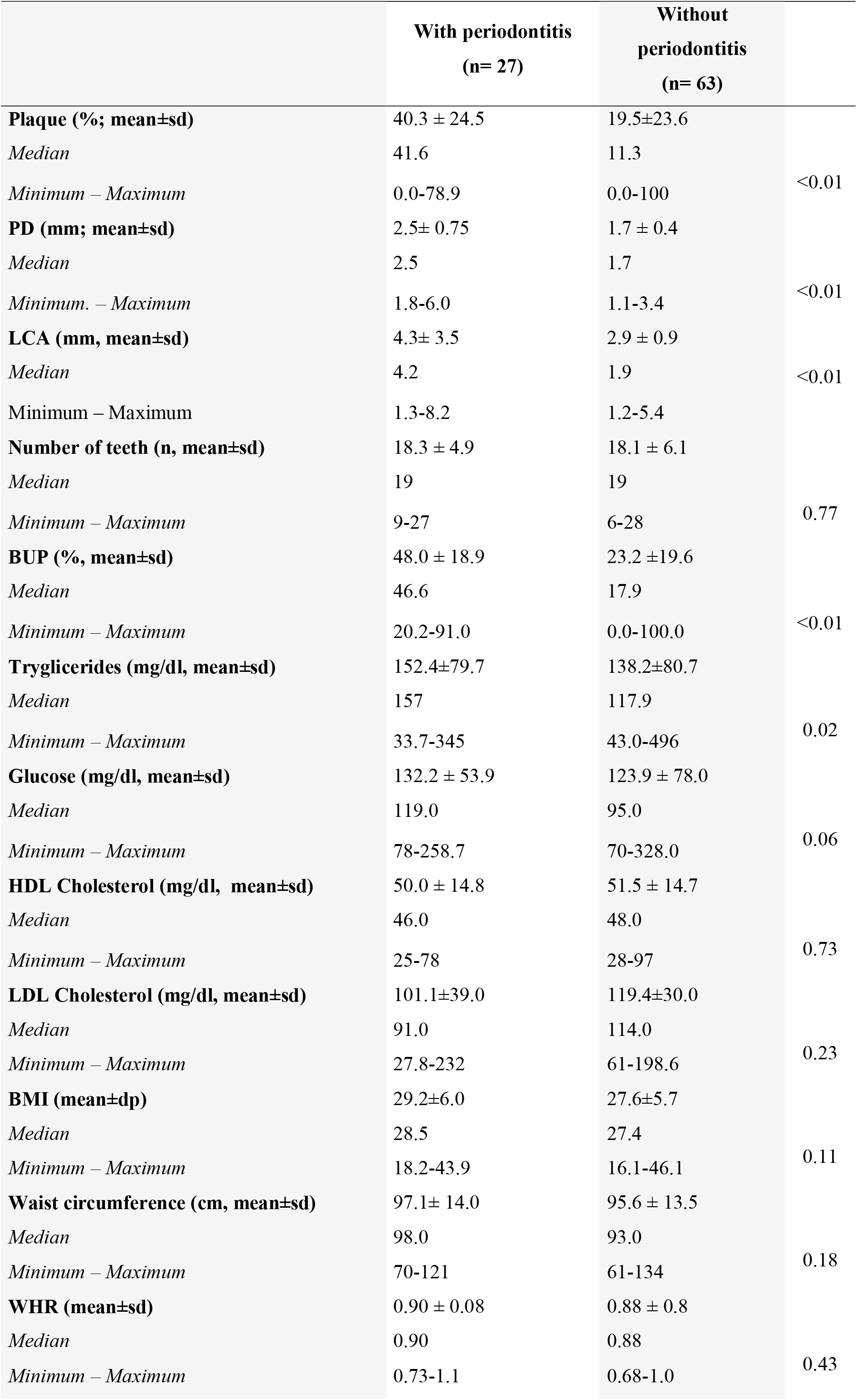
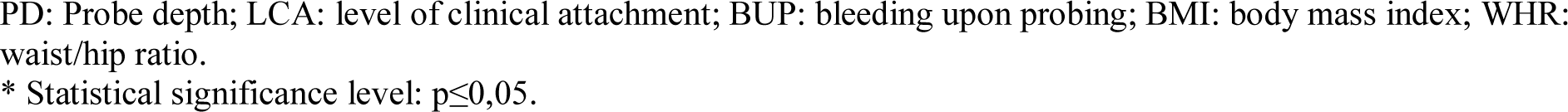
Clinical and laboratorial characteristics and clinical periodontal parameters from study participants, regarding presence or absence of periodontitis, according to periodontitis diagnosis criterion proposed by Gomes-Filho et al (2018). Salvador-Bahia, Brazil, 2018. (n= 90)

|  | With periodontitis<br>(n= 27) | Without<br>periodontitis<br>(n= 63) |  |
| --- | --- | --- | --- |
| **Plaque (%; mean±sd)** | 40.3 ± 24.5 | 19.5±23.6 |  |
| *Median* | 41.6 | 11.3 |  |
| *Minimum – Maximum* | 0.0-78.9 | 0.0-100 | <0.01 |
| **PD (mm; mean±sd)** | 2.5± 0.75 | 1.7 ± 0.4 |  |
| *Median* | 2.5 | 1.7 |  |
| *Minimum. – Maximum* | 1.8-6.0 | 1.1-3.4 | <0.01 |
| **LCA (mm, mean±sd)** | 4.3± 3.5 | 2.9 ± 0.9 |  |
| *Median* | 4.2 | 1.9 | <0.01 |
| *Minimum – Maximum* | 1.3-8.2 | 1.2-5.4 |  |
| **Number of teeth (n, mean±sd)** | 18.3 ± 4.9 | 18.1 ± 6.1 |  |
| *Median* | 19 | 19 |  |
| *Minimum – Maximum* | 9-27 | 6-28 | 0.77 |
| **BUP (%; mean±sd)** | 48.0 ± 18.9 | 23.2 ±19.6 |  |
| *Median* | 46.6 | 17.9 |  |
| *Minimum – Maximum* | 20.2-91.0 | 0.0-100.0 | <0.01 |
| **Tryglicerides (mg/dl, mean±sd)** | 152.4±79.7 | 138.2±80.7 |  |
| *Median* | 157 | 117.9 |  |
| *Minimum – Maximum* | 33.7-345 | 43.0-496 | 0.02 |
| **Glucose (mg/dl, mean±sd)** | 132.2 ± 53.9 | 123.9 ± 78.0 |  |
| *Median* | 119.0 | 95.0 |  |
| *Minimum – Maximum* | 78-258.7 | 70-328.0 | 0.06 |
| **HDL Cholesterol (mg/dl, mean±sd)** | 50.0 ± 14.8 | 51.5 ± 14.7 |  |
| *Median* | 46.0 | 48.0 |  |
| *Minimum – Maximum* | 25-78 | 28-97 | 0.73 |
| **LDL Cholesterol (mg/dl, mean±sd)** | 101.1±39.0 | 119.4±30.0 |  |
| *Median* | 91.0 | 114.0 |  |
| *Minimum – Maximum* | 27.8-232 | 61-198.6 | 0.23 |
| **BMI (mean±dp)** | 29.2±6.0 | 27.6±5.7 |  |
| *Median* | 28.5 | 27.4 |  |
| *Minimum – Maximum* | 18.2-43.9 | 16.1-46.1 | 0.11 |
| **Waist circumference (cm, mean±sd)** | 97.1± 14.0 | 95.6 ± 13.5 |  |
| *Median* | 98.0 | 93.0 |  |
| *Minimum – Maximum* | 70-121 | 61-134 | 0.18 |
| **WHR (mean±sd)** | 0.90 ± 0.08 | 0.88 ± 0.8 |  |
| *Median* | 0.90 | 0.88 |  |
| *Minimum – Maximum* | 0.73-1.1 | 0.68-1.0 | 0.43 |

PD: Probe depth; LCA: level of clinical attachment; BUP: bleeding upon probing; BMI: body mass index; WHR: waist/hip ratio.
\* Statistical significance level: $p \leq 0,05$ .

Finally, we observed a statistical significant association between periodontitis and MetS on both crude and adjusted – by age, family income, alcoholic beverage drinking and physical activity – models (O*R*_***crude***_= 2.58; C I95% [1.02 – 6.55])/ (O*R*_***adjusted***_= 2.63; CI95% [1.01 – 6.80]), and between severe periodontits and MetS (O*R*_***crude***_= 3.86; CI95% [1.24 – 11.98])/ (O*R*_***adjusted***_= 4.14; CI95% [1.29 – 13.29]) (TABLE 4).

**Table 4.** Odds Ratio (OR) and Confidence Interval (CI 95%) obtained by non conditional logistic regression for the associations between periodontitis, according to diagnosis criterion proposed by Gomes-Filho et al. (2018) and MetS, and according to NCEP-ATP III criterion, regarding periodontitis (mild, moderate and severe) vs MetS; moderate and mild periodontitis vs MetS; and severe periodontitis vs MetS. Salvador-Bahia, Brazil, 2018. (n= 90)

| PERIODONTITIS | MODELS | OR | CI 95% | p* |
| --- | --- | --- | --- | --- |
| **Peridontitis** | Crude (n=90) | 2.58 | (1.02 – 6.55) | 0.05 |
|  | Adjusted<sup>1</sup> (n=90) | 2.63 | (1.01 – 6.80) | 0.05 |
| **Moderate e Mild Peridontitis** | Crude (n=72) | 1.17 | (0.27 – 4.90) | 0.79 |
|  | Adjusted <sup>1</sup> (n=72) | 1.16 | (0.28 – 4.90) | 0.83 |
| **Severe Peridontitis** | Crude (n=90) | 3.86 | (1.24- 11.98) | 0.02 |
|  | Adjusted <sup>1</sup> (n=90) | 4.14 | (1.29- 13.29) | 0.02 |

<sup>1</sup> Adjusted by age, family income, alcoholic beverage drinking and physical activity.
\* Statistical significance level: $p \leq 0.05$ .

When estimating ORs on models for the association between periodontitis^36^ and MetS, and regarding criterion of IDF^40^ (TABLE 5), we only observed a significant statistical association between periodontitis and MetS.

**Table 5.** Association between periodontitits (mild, moderate and severe) and MetS, and severe periodontitis and MetS; MetS defined by criterion from NCEP-ATP III and from IDF. Salvador-Bahia, Brazil, 2018. –

| GOMES-FILHO et al., 2018 | Models | NCEP- ATP III |  |  | IDF |  |  |
| --- | --- | --- | --- | --- | --- | --- | --- |
|  |  | OR | CI 95% | p* | OR | CI 95% | p* |
| **Peridontitis** | Crude | 2.58 | (1.02 – 6.55) | 0.05 | 2.78 | (1.07 – 7.30) | 0.04 |
|  | Adjusted | 2.63** | (1.01 – 6.80) | 0.05 | 2.90 | (1.08 – 7.85) | 0.04 |
| **Severe Peridontitis** | Crude | 3.86 | (1.24- 11.98) | 0.02 | 2.74 | (0.89 - 8.51) | 0.08 |
|  | Adjusted | 4.14*** | (1.29- 13.29) | 0.02 | 3.08 | (0.95 - 6.03) | 0.06 |

\* Statistical significance level: $p \leq 0.05$ .
\*\* Adjusted by age, family income, alcoholic beverage drinking and physical activity
\*\*\* Adjusted by age, family income, physical activity and current smoking habit.

When these measures were estimated with periodontitis diagnosis determined by Page and Eke (2007)^37^ and Eke et al. (2012)^38^ criterion, and MetS diagnosis by NCEP-ATP III^39^ criterion and repeated for IDF^40^, we observed that these association only remained significant between periodontitis and MetS (determined by IDF^40^ criterion) (p= 0.04), for both crude and adjusted models (TABLE 06). In order to verify the agreement level among criteria used, Kappa index was applied, which indicated a good agreement between MetS criteria (Kappa= 0.82) and moderate for periodontitis criteria (Kappa= 0.55).

**Table 6.** Association between chronic periodontitis (mild, moderate and severe), severe periodontitis (n= 18) – both determined by Page and Eke (2007) and Eke et al, (2012) criterion - and MetS (by NCEP-ATP III and IDF criteria). Salvador-Bahia, Brazil, 2018. (n= 90)

| **PAGE&EKE, 2007;**<br>**EKE et al.,2012** | **Models** | **NCEP-ATP III** |  |  | **IDF** |  |  |
| --- | --- | --- | --- | --- | --- | --- | --- |
|  |  | **OR** | **CI 95%** | **p\*** | **OR** | **CI 95%** | **p\*** |
| **Periodontitis** | Crude | 2.55 | (0.96 – 6,70) | 0.06 | 2.95 | (1.13 – 7.63) | 0.04 |
|  | Adjusted<sup>1</sup> | 2.49 | (0.88 – 7.06) | 0.12 | 3.21 | (1.13 – 9.12) | 0.04 |
| **Severe periodontitis** | Crude | 2.10 | (0.84- 4.79) | 0.12 | 2.38 | (0. 89 - 5.81) | 0.06 |
|  | Adjusted<sup>1</sup> | 4.14 | (0.86- 5.15) | 0.11 | 2.40 | (0.95 - 6.03) | 0.06 |

<sup>1</sup> Adjusted by age, family income, current smoking habit and physical activity
\* Statistical significance level: $p \leq 0.05$ .

## DISCUSSION

Preliminary results observed in the present study indicate an association between periodontitis (Gomes-Filho et al., 2018)^36^ and Metabolic Syndrome (NCEP-ATP III)^39^, related to the presence of both periodontitis and severe periodontitis and when estimated in the model adjusted by confounding covariables: age, family income, alcoholic beverage drinking and physical activity. Studies from different countries also observed this association between MetS and periodontitis ^20,31,32,41^, other studies only observed it in women ^42,43^, while one study found this association only in men ^44^.

The bigger effect of this association regarding only severe periodontitis supports studies that only observed this association. Biological plausibility for the inter-relationship between periodontitis and MetS seems to be based by evidence showing the role of immunological markers in serum levels, such as IL-6 and TNF, that interconnect both diseases, in which combined effects have a synergic role on the coexistence of periodontitis and MetS^19,20,26,44–48^.

We did not observe an association between severe periodontitis, according to Gomes Filho et al.(2018)^36^ criterion, and MetS (IDF criterion)^40^. Despite the good agreement verified between both criteria for the diagnosis of MetS in the sample, this divergence in the finding of association of severe periodontitis reveals that the disagreements that persists in relation to the referred association, may be due to issues of this type^47^.

There was also a difference on the association between periodontitis, according to Page and Eke (2007)^37^ and Eke et al (2012)^38^ and MetS – by both NCEP-ATP III^40^ and IDF diagnosis criteria^40^, showing an association only for periodontitis and MetS (IDF criterion) on the adjusted model. These differences show the lack of standardizing of diagnosis criteria for periodontitis, which may be related to the existence of different evidence regarding the association of periodontitis and MetS. Furthermore, the diagnosis criterion chosen may cause bias on association measurement and compromise the quality of this measure^49^.

The prevalence of MetS was 46.67%, which is similar to other Brazilian studies. Aragão, Bós and Souza (2014)^50^ estimated a MetS prevalence of 55.4% in a quilombola community of Piauí state, and Borges (2007)^18^, that evaluated Japanese-Brazilian participants observed a prevalence of 54.3%. Studies performed in rural populations found lower estimates of MetS prevalence. Oliveira, Souza and Lima (2006)^51^, in an assessment of a rural population in semiarid region of Bahia, showed a 30% prevalence of MetS. Haab, Benvegnú and Fisher (2012)^52^ found a 15.6% MetS prevalence in a rural community of Santa Rosa County in the state of Santa Catarina. These differences indicate an influence of socio-cultural and population characteristics on MetS prevalence estimates.

Regarding population assisted in health units in urban regions, the prevalence of MetS estimated in the present study is similar to the outcomes observed by Leitão and Martins (2012)^53^, for instance, a 56.7% MetS prevalence in Sao Paulo regions with low socioeconomic indexes, and 34.0% in those with higher socioeconomic indexes. The higher prevalence observed in lower socioeconomic status regions points to poverty as a risk factor for MetS^53^.

Metabolic Syndrome is associated with diabetes due to an increase in insulin resistance in individuals with MetS,^9^ which results in a higher number of cardio metabolic factors^54^. Thus, the results observed in a meta-analysis performed by Liu et al. (2019)^55^ were weak and/or inconclusive regarding the potential effect of periodontal therapy on the prevention of cardiovascular diseases, and the meta-analysis from Baeza et al. (2020)^56^ also indicated inconclusive evidences for the effect of periodontal therapy on glucose control.

According to Souza et al. (2015)^57^, MetS and its components were significantly more common in obese individuals. The role of obesity in MetS is due to the increase of insulin resistance indexes, for instance, the gradual increase of obesity and changes in plasma metabolites, such as increases in total cholesterol and triglycerides, resulting from obesity^58^.

The prevalence of periodontitis observed in the present study was higher than the national average, which applyied the Community Periodontal Index (CPI) as a diagnosis criterion, where the prevalence of moderate to severe periodontitis was 15.8%, and severe periodontitis only 5.8%^59^. This difference is due to different criteria used for the diagnosis of periodontitis, where the use of CPI may underestimate the disease prevalence, due to the use of índex teeth and partial examination^60^.

This outcome is confirmed by the elevated loss of LCA, which was significantly higher in those diagnosed with periodontitis. Similar findings were observed by Borges (2007)^18^ that associated LCA loss to dental loss due to periodontal diseases. It is important to emphasize that Adachi and Kobayashi (2020)^61^ observed an effect independent of dental loss on MetS.

There was a positive association between periodontitis and serum triglyceride levels. Loscheet et al. (2000)^62^ also observed a significant increase in these levels in individuals with periodontitis. This seems to be associated to cytokine clearance in response to infection by gram-negative bacteria, where hyperlipidemia may occur due to these chemical mediators^63^.

This preliminary study has limitations, such as the insufficient sample size and its convenience selection method. Initially, it limits the study power, influencing on outcome significance and restricting the generalization of results. Moreover, this is a cross-sectional study, which does not allow inferences on cause-effect relationship between periodontitis and MetS, although they are interconnected.

## CONCLUSIONS

The main results from the present study indicate the positive association between periodontitis and metabolic syndrome, where there was observed a higher effect when associated only to severe periodontitis cases. These results enhance the significant association between periodontitis and MetS shown in literature, especially regarding periodontitis severity. The variety of findings and oppositions that are still present in the literature, however, show that this association is not yet well determined, showing a need for more studies that show this association. Finally, it is unquestionable the role of oral teams on individuals health, especially public, raising a professional profile more integrated with the promotion of periodontal health and overall wellbeing of health services users.

## Data Availability

There is no support from any organization for the submitted work; no financial relationships with any organizations that may have an interest in the work presented in the previous three years; no other relationships or activities that may appear to have influenced the submitted work.

